# Machine learning with validation to detect diabetic microvascular complications using clinical and metabolomics data

**DOI:** 10.1101/2022.08.12.22278659

**Authors:** Feng He, Clarissa Ng Yin Ling, Simon Nusinovici, Ching-Yu Cheng, Tien Y. Wong, Jialiang Li, Charumathi Sabanayagam

## Abstract

**AIMS:** Using machine learning integrated with clinical and metabolomic data to identify biomarkers associated with diabetic kidney disease (DKD) and diabetic retinopathy (DR), and to improve the performance of DKD/DR detection models beyond traditional risk factors.

**METHODS:** We examined a population-based cross-sectional sample of 2,772 adults with type 1 or type 2 diabetes from Singapore Epidemiology of Eye Diseases study (SEED, 2004-2011). LASSO logistic regression (LASSO) and gradient boosting decision tree (GBDT) were used to select markers of prevalent DKD (defined as an eGFR < 60ml/min/1.73m2) and prevalent DR (defined as an ETDRS severity level ≥ 20) from an expanded set of 19 established risk factors and 220 NMR-quantified circulating metabolites. Risk assessment models were developed based on the variable selection results and externally validated in UK Biobank (n=5,843, 2007-2010). Model performance (AUC with 95% CI, sensitivity, and specificity) of machine learning was compared to that of traditional logistic regression adjusted for age, gender, diabetes duration, HbA_1c_%, systolic BP, and BMI.

**RESULTS:** SEED participants had a median age of 61.7 years, with 49.1% female, 20.2% having DKD, and 25.4% having DR. UK Biobank participants had a median age of 61.0 years, with 39.2% female, 6.4% having DKD, and 5.7% having DR. Both algorithms identified diabetes duration, insulin usage, age, and tyrosine as the most important factors of both DKD and DR. DKD was additionally associated with CVD, hypertension medication, and three metabolites (lactate, citrate, and cholesterol esters to total lipids ratio in intermediate-density-lipoprotein); While DR was additionally associated with HbA_1c_, blood glucose, pulse pressure, and alanine. Machine-learned models for DKD and DR detection outperformed traditional logistic regression in both internal (AUC: 0.832-0.838 vs. 0.743 for DKD, and 0.779-0.790 vs. 0.764 for DR) and external validation (AUC: 0.737-0.790 vs. 0.692 for DKD, and 0.778 vs. 0.760 for DR).

**CONCLUSIONS:** Machine-learned biomarkers suggested insulin resistance to be a primary factor associated with diabetic microvascular complications. Integrating machine learning with biomedical big data enabled biomarker discovery from a wide range of correlated variables, which may facilitate our understanding of the disease mechanisms and improve disease screening.

## INTRODUCTION

Diabetes is one of the most prevalent and serious health problems of our times. International Diabetes Federation estimated the global adult population with diabetes to be 536.6 million in 2021, and projected it to reach 783.2 million by 2045 [1]. With this rapid-growing population and the greater longevity over time, the burden of diabetic complications is expected to increase in parallel [2, 3].

Diabetic kidney disease (DKD) and diabetic retinopathy (DR) are diabetic microvascular complications known to decrease quality of life, cause disability or even premature death if undetected and untreated [4, 5]. However, timely and accurate diagnosis remains a challenge for those at risk because of the asymptomatic progression in early stages [6]. Age, gender, diabetes duration, HbA_1c_, systolic BP, and BMI have been identified as the major risk factors [7], yet they do not fully account for the variation in risk faced by different individuals. Evidence showed a connection between DR and DKD, suggesting some shared pathogenic pathways, or both being manifestations of a latent systematic microvasculature disease [3, 8]. However, previous studies in search of useful biomarkers were often hampered by inadequate data availability, lack of replication, and limited data analysis methods, unable to examined a wide range of variables simultaneously [4, 6]. As a promising solution, machine learning integrated with biomedical big data has been implemented for biomarker discovery for DKD and DR individually [9, 10]. However, to the best of our knowledge, few studies have used machine learning to investigate the commonalities and differences of these two tissue-specific complications in terms of metabolomic profiling, which may serve as a window to reveal the latent biochemical changes and hidden pathogenic pathways [6].

Herein we aim to fill these gaps by implementing two classic machine learning algorithms - LASSO logistic regression (LASSO) and gradient boosting decision tree (GBDT), to simultaneously examine 239 variables (19 established risk factors and 220 circulating metabolites) as predictors of prevalent DKD/DR in a retrospective Asian adult cohort with type 1 or type 2 diabetes. Risk assessment models were developed based on machine-learned biomarkers, and externally validated using UK Biobank data, against the traditional logistic regression in terms of AUC (95% CI), sensitivity, and specificity.

## MATERIALS AND METHODS

### Datasets and inclusion/exclusion criteria

We derived the study data from Singapore Epidemiology of Eye Diseases study (SEED), a population-based cross-sectional study conducted in Singapore from 2004 to 2011, with methodological details reported elsewhere [11]. In brief, we recruited participants aged 40-80 years in an age-stratified random sampling manner and asked them to take the interviewer-administered questionnaire, ocular examinations, and biochemical laboratory tests. 10,033 adults were successfully recruited, including 3,280 Malays (2004–2006, response rate 78.7%), 3,400 Indians (2007–2009, 75.6%), and 3,353 Chinese (2009–2011, 72.8%). Of these, we excluded participants free of diabetes (*n*=7,069), which was defined as having an HbA_1c_% > 6.5, random blood glucose > 11.1 mmol/L, self-reported physician-diagnosed diabetes, or the use of anti-diabetic medication including insulin. We also excluded those missing metabolomics profiles (*n*=179), or missing more than 10% of the data (*n*=13), to get a final study population of 2,772 individuals (**Figure 1**).

**Figure 1.**
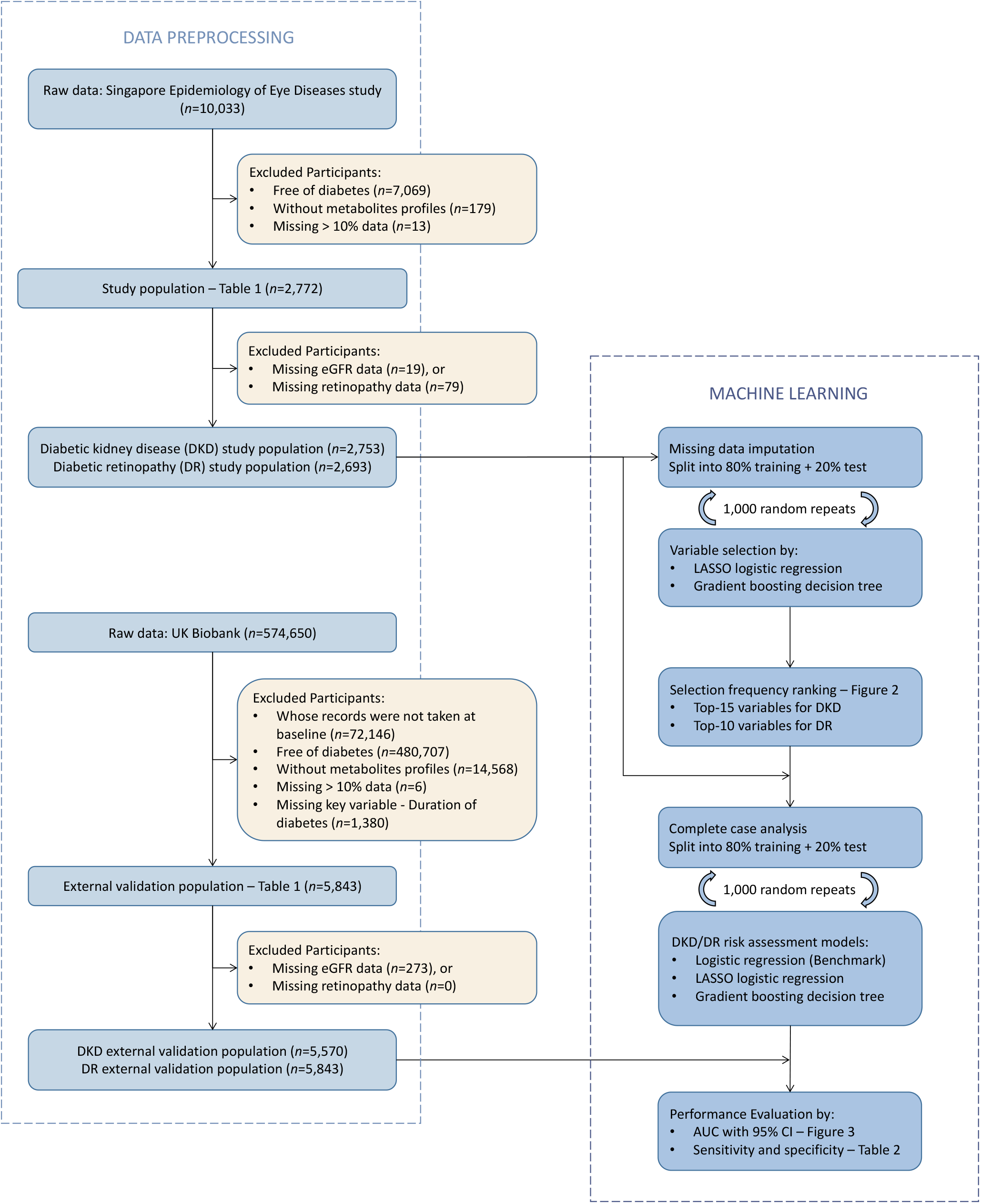
Analysis flow diagram.

For external validation, we extracted data from UK Biobank (UKBB), an open access resource of prospective dataset collected in the United Kingdom from 2007 to 2010, with over 500,000 participants [12]. Diabetes was defined the same as that of SEED, but additionally included those with DR if the aforementioned variables were not available. After data pre-processing, 5,843 participants were found eligible for the external validation.

Both SEED and UKBB were conducted in accordance to the Declaration of Helsinki, with the ethics approval obtained from SingHealth Institutional Review Board and the North West Multi-Centre Research Ethics Committee, respectively. Written informed consent was provided by all participants.

### Outcomes and covariates

DKD was defined as an eGFR<60 ml/min/1.73m2 for both SEED and UKBB in participants with diabetes, where the eGFR values were calculated from blood creatinine concentrations using the chronic kidney disease epidemiology collaboration (CKD-EPI) equation [13]. DR severity in each eye of SEED participants was graded from fundus photographs by certified ophthalmic graders according to the standard protocol of Early Treatment Diabetic Retinopathy Study (ETDRS) [2, 14]. Based on ETDRS severity score levels, DR severity was categorized into 5 stages: minimal (level 20), mild (level 35), moderate (levels 43 to 47), severe (level 53), and proliferative DR (levels > 60). For the current study, the outcome was “any DR” defined as an ETDRS level ≥ 20 in at least one eye. We also defined an alternative outcome in supplementary analysis – moderate and above DR (level > 43 in at least one eye). In UKBB, DR severity was not graded, therefore, we defined “any DR” as those having ICD-10 code “H36.0” in their health-related outcomes (Data-Field: 41270) [12].

For variable selection, we included 239 variables (**Table S1**). Of these, 19 variables were identified by literature review, including 6 traditional risk factors (age, gender, duration of diabetes, HbA_1c_%, systolic BP, and BMI), and 13 extended risk factors related to lifestyle (alcohol drink and smoking), medication use (insulin, anti-cholesterol, and anti-hypertensive medication), clinic/biochemistry (diastolic BP, pulse pressure (PP), random blood glucose, cholesterol, HDL cholesterol, and LDL cholesterol), and comorbidity conditions (CVD and hypertension). Hypertension in both cohorts was manually defined as self-reported physician-diagnosed hypertension, systolic BP > 140 mmHg and diastolic BP > 80 mmHg, or the use of antihypertensive medication. Using NMR techniques (Nightingale Health, Helsinki, Finland), we quantified the concentration of 228 circulating metabolites from patients’ blood samples. Of these, glycerol, pyruvate, and glutamine were not available for Malays; Creatinine was used in eGFR calculation and DKD outcome definition; While four metabolites (total, HDL, and LDL cholesterols, and random blood glucose) were duplicated with those measured in biochemistry tests. Hence for the current study, we only included the remaining 220 metabolites from 15 categories (amino acids, apolipoproteins, cholesterol, cholesterol esters, fatty acids, fluid balance, free cholesterols, glycolysis related metabolites, inflammation, ketone bodies, triglycerides, lipoprotein particle sizes, lipoprotein subclasses, lipoprotein lipid ratios, and other lipids).

### Machine learning algorithms

We used logistic regression with the least absolute square shrinkage operator (LASSO [15]) and gradient boosting decision tree (GBDT [16]) to derive and validate the risk assessment models of DR and DKD. LASSO is an extension of traditional logistic regression (LR) that does not require the independence of covariates. Therefore, this algorithm is often used in high-dimensional setting where multi-collinearity could be an issue. During the course of parameter optimization, LASSO automatically shrinks the coefficients of those less-important variables to zeros, while leaving those important variables with non-zero coefficients to achieve biomarker selection. Its strength was being relatively simple in computation complexity and parameter tuning as compared to other machine learning algorithms, but is limited to examining only the linear associations between continuous covariates and the log-odds. To account for possible non-linear terms and variable interactions, we additionally implemented GBDT, which essentially is a sequence of inter-dependent decision tree models. This algorithm is well-known for its adaptability to various data distributions in prediction and variable selection tasks. However, it is rather time-consuming in terms of hyper-parameter tuning and computation, also more likely to overfit data, and less transparent as compared to LASSO.

### Statistical analysis

We conducted all analyses in R 4.0.2., and defined statistical significance as a p-value < 0.05. We described the population characteristics of SEED and UKBB using *n* (%), mean (SD), or median [IQR] as appropriate for the variable (**Table 1**). Some sub-categories may not add up due to the presence of missing data. Pearson’s Chi-square tests and Mann-Whitney “U” tests were used to compare characteristics of the two study populations, which indicated significant difference (p<0.001) in all aspects compared.

**Table 1.** Comparison of SEED and UKBB population characteristics.

|  | **SEED (n=2772)** | **UK Biobank (n=5843)** |
| --- | --- | --- |
| Age, years | 61.7 [53.5, 69.4] | 61.0 [55.0, 65.0] |
| Gender = Female | 1361 (49.1) | 2090 (35.8) |
| Ethnicity | Malay, Indian, Chinese | British, Irish, African, etc. |
| CKD, % | 555 (20.2) | 377 (6.8) |
| eGFR, ml/min/1.73m<sup>2</sup> | 78.9 (23.0) | 88.6 (17.1) |
| Any DR, % | 685 (25.4) | 355 (6.1) |
| Duration of diabetes, years | 3.9 [0.0, 10.7] | 5.0 [2.0, 10.0] |
| HbA<sub>1c</sub>, % | 7.7 (1.7) | 7.0 (1.3) |
| Blood glucose, mmol/L | 9.8 (4.8) | 7.6 (3.4) |
| Systolic BP, mm Hg | 145.5 (22.2) | 142.9 (18.2) |
| Diastolic BP, mm Hg | 78.3 (10.5) | 81.2 (10.4) |
| Pulse pressure, mm Hg | 67.1 (18.0) | 74.3 (13.3) |
| BMI, Kg/m<sup>2</sup> | 26.9 (4.8) | 31.3 (5.7) |
| Total cholesterol, mmol/L | 5.2 (1.2) | 4.5 (1.0) |
| HDL cholesterol, mmol/L | 1.1 (0.3) | 1.2 (0.3) |
| LDL cholesterol, mmol/L | 3.2 (1.0) | 2.7 (0.7) |
| Insulin = Yes | 143 (5.2) | 1245 (21.3) |
| Anti-cholesterol medication = Yes | 1183 (43.6) | 4479 (76.7) |
| Anti-hypertensive medication = Yes | 1474 (53.4) | 3727 (63.8) |
| CVD = Yes | 519 (18.7) | 245 (4.2) |
| Hypertension = Yes | 2228 (80.5) | 4164 (71.3) |
| Current smoker = Yes | 377 (13.6) | 3193 (54.6) |
| Alcohol consumption = Yes | 204 (7.4) | 5342 (91.4) |

\*Data presented as count (%), mean (SD) or median [IQR] as appropriate for the variable

In SEED, the missing proportions were controlled below 10% for each variable and below 6% for each participant. We assumed data missing at random and performed missing data imputation using mean values/modes as appropriate for each variable to maximize the sample size for variable selection. To reduce selection bias caused by training and test set split, we averaged the results across 200 random repeats of five-fold cross-validation. In each repeat, the imputed SEED dataset was randomly divided into 5 subsets (i.e., folds) of equal sample size and case rate by stratified sampling. Each fold (20%) took turns to be the validation set, while the remaining four (80% data) were used for model training and variable selection. From 200 replicates we generated 1,000 sets of selected variables, based on which we quantified the contribution of each variable to the model performance as a variable importance score, calculated the variable selection frequency (%) during the repeated cross-validation.

Next, we ranked the variables according to their selection frequencies from high to low (**Figure 2**), and took the top-15 associated with DKD, and the top-10 associated with DR, respectively, to derive the disease risk assessment models using the same two machine learning algorithms. To evaluate the performance of these new models, we performed another 200 random repeats of 5-fold cross-validation but used only the complete cases (i.e., no missing data imputation). As a performance reference, we developed logistic regression (LR) models adjusted for the 6 traditional risk factors. The model performance metrics included the AUC with 95% CI (**Figure 3**), sensitivity at 70% specificity, and specificity at 80% sensitivity (**Table 2**).

**Table 2.** Machine Learning Model Performance*.

|  |  | Sensitivity at<br>70% specificity | Specificity at<br>80% sensitivity | *n* | Number of cases<br>(%) |
| --- | --- | --- | --- | --- | --- |
| *Diabetic Kidney Disease* |  |  |  |  |  |
| SEED | LR | 64.9 | 54.0 | 2,653 | 517 (19.5) |
|  | LASSO | 80.0 | 70.3 | 2,666 | 532 (20.0) |
|  | GBDT | 80.4 | 70.9 | 2,668 | 529 (19.8) |
| UKBB | LR | 56.4 | 47.2 | 5,236 | 348 (6.6) |
|  | LASSO | 74.5 | 62.8 | 5,089 | 336 (6.6) |
|  | GBDT | 62.5 | 53.2 | 5,543 | 369 (6.7) |
| *Diabetic Retinopathy* |  |  |  |  |  |
| SEED | LR | 69.1 | 59.4 | 2,597 | 653 (25.1) |
|  | LASSO | 71.7 | 59.6 | 2,514 | 628 (25.0) |
|  | GBDT | 71.9 | 61.6 | 2,598 | 655 (25.2) |
| UKBB | LR | 71.1 | 57.1 | 5,492 | 336 (6.1) |
|  | LASSO | 73.3 | 61.7 | 4,678 | 280 (6.0) |
|  | GBDT | 72.8 | 60.4 | 4,833 | 296 (6.1) |

\* Results from complete case analysis (i.e., no missing values). Sensitivity and specificity scores were averaged over 1,000 replicates from 200 random repeats of 5-fold cross validation.
LR models were adjusted for 6 traditional risk factors (age, gender, HbA<sub>1c</sub>%, systolic BP, BMI, and duration of diabetes). GBDT and LASSO models for DKD included top-15 variables, whereas the models for DR included top-10.

**Figure 2.**
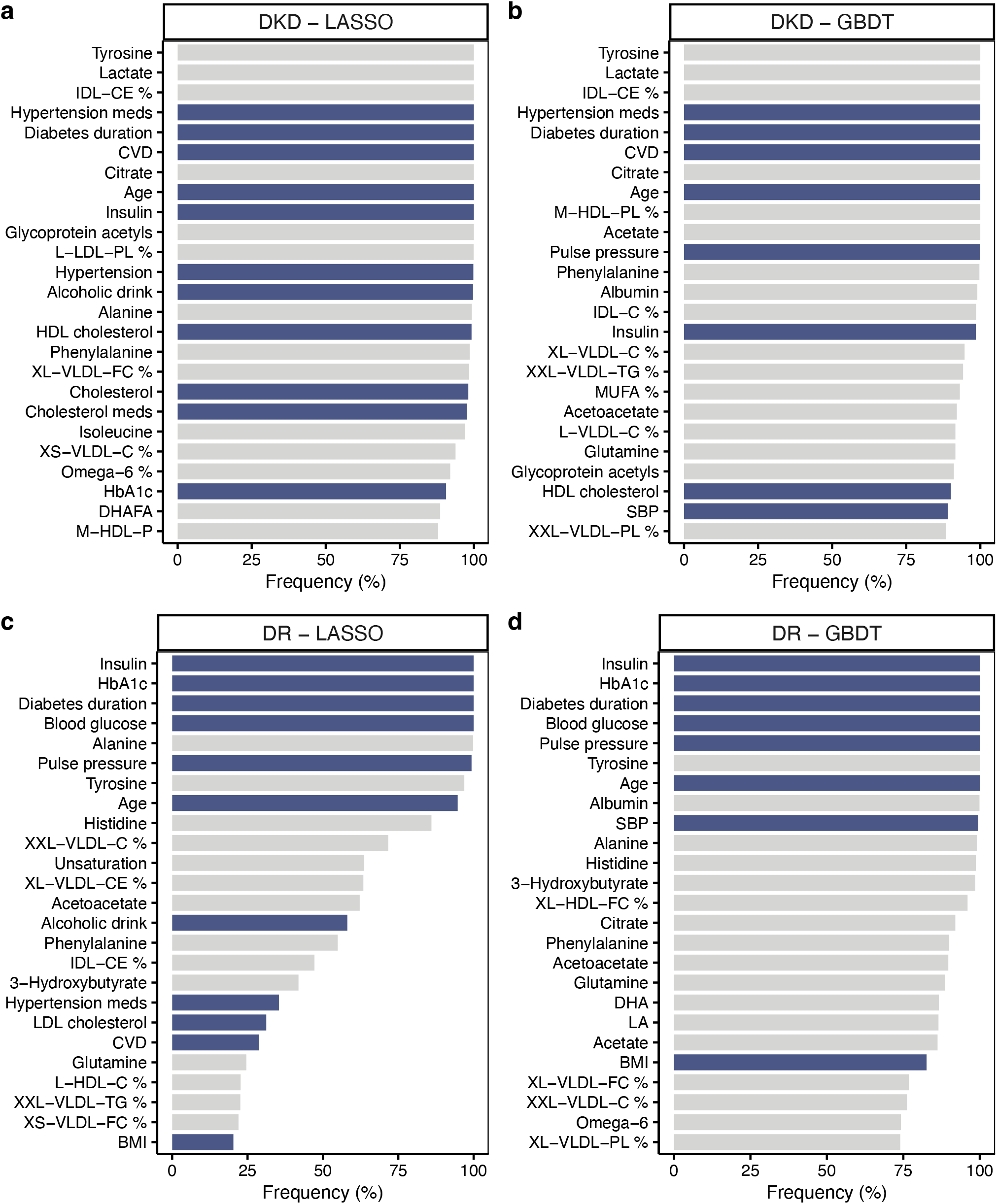
Bar plots showing the top-25 variables selected by machine learning. HDL: high-density lipoprotein; IDL: intermediate-density lipoprotein; LDL: low-density lipoprotein; VLDL: very-low-density lipoprotein;L: large; M: medium; S: small; XL: very large, XS: very small; XXL: extremely large; D: mean diameter; C: cholesterol; CE: cholesterol esters; FC: free cholesterol; L: total lipids; PL: phospholipids; TG: triglycerides; %: ratio to total lipids; FA: fatty acids;

**Figure 3.**
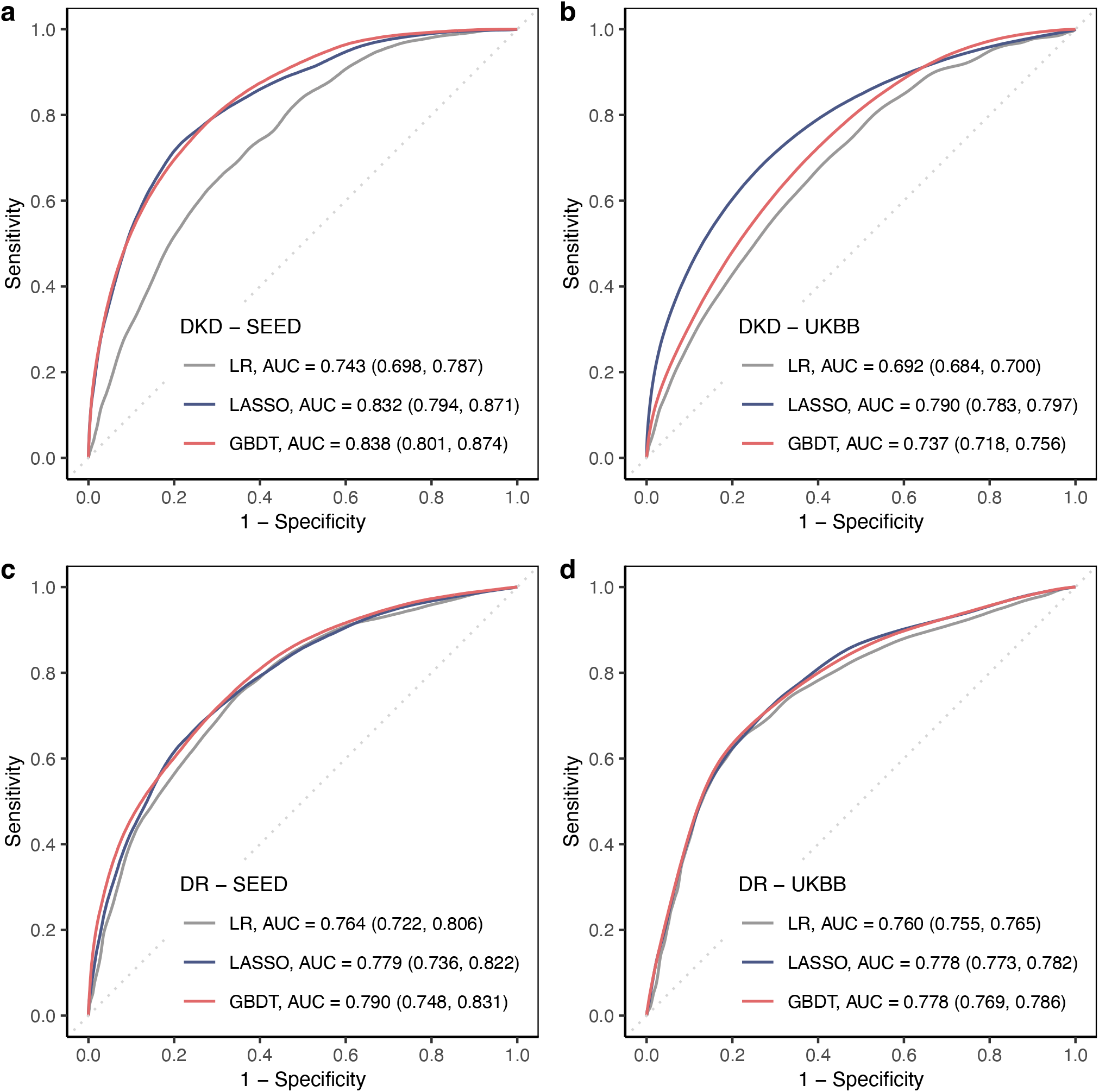
Receiver operating characteristic curves. LR adjusted for 6 traditional factors (age, gender, HbA_1c_%, systolic BP, BMI, and duration of diabetes). For DKD detection, both GBDT and LASSO used the corresponding top-15 variables, whereas for DR they both used top-10.

## RESULTS

### Population characteristics

SEED diabetic population included 2,772 individuals, with a median age of 61.7 [53.5, 69.4] years and 49.1% being female. UKBB diabetic population included 5,843 individuals, with a median age of 61.0 [55.0, 65.0] years and 35.8% being female. SEED participants showed a higher prevalence of both DKD and DR as compared to the UKBB participants (DKD 20.2% vs. 6.8%, and DR 25.4% vs. 6.1%). Moreover, around 6.6% of the SEED population developed both DKD and DR, while in UKBB, only 0.7% were found with both complications. The two study populations also differed significantly in terms of lifestyle, demographic factors, lab results, and comorbidities (**Table 1**, all p-values < 0.001, data not shown).

### Variable importance ranking

The selection frequency of 239 variables were used as an indicator of variable importance (**Figure 2**). For DKD, the top-8 variables selected by LASSO and GBDT were the same, with 100% selection frequency, including four traditional risk factors (duration of diabetes, CVD, hypertensive medication use, and age) and four metabolites (tyrosine, lactate, cholesterol esters to total lipid ratios in intermediate-density-lipoprotein [IDL-CE%], and citrate). The use of insulin was also selected by both but with slightly lower frequency by GBDT for DKD (=98.5%). For any DR, only the top-4 variables selected by the two algorithms were the same, including insulin use, HbA_1c_, duration of diabetes, and blood glucose, all with 100% frequency. Additionally, age, PP and two metabolites (tyrosine and alanine) were found important for DR. For moderate and above DR, the same 6 risk factors and tyrosine were selected as top variables (**Supplementary Figure S3, and S4**). Based on **Figure 2**., we decided to use the top-15 variables for the development of DKD screening models, and the top-10 for DR.

### Model performance

ROC curves in internal and external validation were shown in **Figure 3**. For DKD, LASSO and GBDT achieved similar performance improvement for SEED participants (AUC = 0.832 by LASSO, 0.838 by GBDT, vs. 0.743 by LR), but for UKBB, LASSO performed significantly better (0.790 by LASSO, vs. 0.737 by GBDT, and 0.692 by LR). For DR, internal validation only showed insignificant differences, yet in external validation, machine learning was again significantly better (0.778 by LASSO, 0.778 by GBDT, vs. 0.760 by LR). We further compared the models in terms of sensitivity and specificity, and found GBDT being the best in internal validation – at 80% sensitivity, it achieved 70.9% specificity for DKD, and 61.6% for DR. In external validation, however, LASSO was the best with specificity 62.8% for DKD and 61.7% for DR (**Table 2**).

## DISCUSSION

### Principal findings

Duration of diabetes, age, use of insulin, and circulating tyrosine were the most important markers for DKD and DR detection in SEED diabetic population. DKD was also associated with the use of antihypertensive medications, CVD, and three metabolites (lactate, citrate, and IDL-CE%); whereas DR was additionally linked to HbA_1c_, random blood glucose, PP, and alanine. Machine learning models outperformed the traditional LR in terms of AUC, sensitivity, and specificity, demonstrating their potential to discover novel biomarkers and enable disease screening when integrated with healthcare and metabolomics data.

### Strengths and limitations

Our main dataset included a comprehensive set of 19 risk factors and 220 circulating metabolites measured in 2,772 individuals. The detailed patient profiling with adequate sample size allowed an opportunity to identify markers most relevant to DKD and DR, offering insights into the systematic alteration of metabolism and underlying pathogenic pathways. Such findings may facilitate novel treatment therapies for those at high risk because metabolites like tyrosine could be manually modulated via dietary intake. For biomarker discovery, traditional studies often rely on logistic regression models to examine metabolites one by one separately [17, 18], with stringent model assumptions and multiple testing correction [19]. Herein machine learning provided a simpler approach to simultaneously examine all variables for potential associations. Although LASSO was limited to detect only the linear associations, we had GBDT as a complementary to additionally evaluate the non-linear terms and complex interactions. As was shown in **Supplementary Fig S1**, metabolites in GBDT had higher selection frequencies than in LASSO, demonstrating the existence of such high-order associations in the circulating metabolite network. Still, external validation found LASSO models with the best performance, indicating a prominent contribution of linear associations to DKD/DR detection. Another highlight of our study was using repeated cross-validation to ensure the randomness of sampling data, thereby generating results more robust than those based on a fixed training set. Repeated cross-validation also allowed us to easily compare the variable relative importance based on their selection frequencies, especially of those highly correlated variables. For instance, our models selected PP with a higher frequency than systolic BP for DR, implying the former to be more predictive of the disease outcome. To further increase the validity, we tested our models in 5,843 samples from UK Biobank, an independent study cohort with significantly different population characteristics from SEED. Results were consistent in that machine learning models based SEED still outperformed logistic regression in terms of AUC, sensitivity, and specificity.

One limitation of the current study was that we did not separate study subjects by diabetes type. Since over 95% of the SEED participants had type 2 diabetes, the variable selection results would mainly reflect their associations with type 2 diabetes. Another issue was data availability – many SEED participants did not have data for albuminuria, an important indicator of kidney disease [4], and three metabolites (pyruvate, glycerol, and glutamine). Hence we did not include these variables for selection. UKBB did not provide ETDRS DR severity information needed to define moderate/above DR, hence we could not validate the supplementary models in UKBB. Finally, it is important to note that our results from a cross-sectional study could at best imply correlations and not causations.

### Implications of this study

Major insights were gained through the evaluation and comparison of the 19 established risk factors and the 220 circulating metabolites (**Supplementary Figure S1 and S2**). For both disease outcomes, different machine learning algorithms identified the same three factors (diabetes duration, age, and the use of insulin), supporting the current consensus on DR and DKD risk factors [4, 20]. Moreover, we noted that a circulating metabolite, tyrosine, was also selected by machine learning with top frequency. This semi-essential amino acid can only be synthesized by the hydroxylation of an essential amino acid called phenylalanine, or supplied via nutritional intake [21]. In people with chronic renal failure, however, reduced phenylalanine hydroxylase activity may indicate impaired kidney function, known to increase the systematic risk of microvascular diseases [18, 21]. Tyrosine is important for molecular recognition mediating [22], and its increased level has been linked to insulin resistance and high diabetes risk in several populations [23, 24]. In SEED, the selection of tyrosine, along with insulin, age and diabetes duration, may indicate prolonged insulin resistance as a primary risk factor of diabetic microvascular complications such as DKD [25].

For DKD prevalence, we additionally found CVD of high importance, pointing to the well-known association between CVD and DKD [26]. The selection of hypertension, PP, and antihypertensive medications underscored the importance of BP control to prevent and postpone disease progression [27]. Of the DKD-specific metabolites, IDL-CE% highlighted the change in IDL composition, pointing to the impaired kidney function for lipoprotein metabolism[28]; Higher citrate level has been found associated with the dysregulation of mitochondrial function in DKD [29]; while lactate metabolism in the kidney cortex, a crucial process for energy production and glucose formation for systemic and medullary use, may be affected by the use of diabetic medication [30, 31]. As citrate and lactate are both glycolysis-related metabolites, their selection may imply changes in glycolysis during the course of DKD, which has been linked to impaired adaptive responses to hypoxia, known to increase diabetic complication risks [32].

Among the DR-specific factors, three out of the top-4 (insulin, HbA_1c_, random blood glucose, and diabetes duration) were directly related to glycaemic control, highlighting the possible glucose intolerance and hyperglycaemia in those at high risk of DR [25]. Of the circulating metabolites, alanine was selected as the DR-specific metabolite. This amino acid plays a key role in gluconeogenesis, and its increased concentration in plasma has been linked to the glucose intolerance and insulin resistance in obesity [33].

Based on variable selection frequencies, we also gained novel insights into the established risk factors of high correlation (**Supplementary Figure S2**). Of the three correlated metrics of blood pressure levels, PP had a higher frequency than systolic BP and diastolic BP in DR models, agreeing with Yamamoto, M., et al. that PP is a better predictor of severe DR incidence than systolic BP [34]. Their hypothesis was that PP as a surrogate marker of arterial stiffness, reflected not only the elevated systolic BP but also reduced diastolic BP, thereby carrying more predictive information of DR than other metrics. However, in DKD models based on GBDT, we did not observe a similar trend. Another pair of correlated indexes were HbA_1c_ and random blood glucose for glycaemia control, of which HbA_1c_ got higher frequency. This was probably because random blood glucose data contained more noises from life cycle changes and inter-individual variability than HbA_1c_. Interestingly, some well-established risk factors usually included such as gender disappeared from the top-ranking lists by machine learning, although this variable had been selected by traditional LR models on the same population in previous studies [35]. This could be because gender is an intrinsic component of other phenotypes. For instance, male gender was associated with CVD [36], well-known to be linked to DR [8, 37]. In DCCT/EDIC study, gender difference was also linked to the association between lipoproteins and DKD [38].

## CONCLUSIONS

Current machine learning study in SEED diabetic population showed age, insulin, diabetes duration, and tyrosine of the highest importance for both DKD and DR detection. Integrating machine learning with biomedical big data allowed biomarker discovery from a wide range of correlated variables, which may facilitate our understanding of the disease and enable disease screening.

## Data Availability

SEED data are available from the Singapore Eye Research Institutional Ethics Committee for researchers who meet the criteria for access to confidential data. Interested researchers can send data access requests to the Singapore Eye Research Institute using the following email address.
The UK Biobank datasets can be requested by bona fide researchers for approved projects, including replication, through HTTPS://WWW.UKBIOBANK.AC.UK/.

HTTPS://WWW.UKBIOBANK.AC.UK/

## ABBREVIATIONS

DKD: Diabetic kidney disease
DR: Diabetic retinopathy
EDIC: Epidemiology of Diabetes Interventions and Complications
ETDRS: Early Treatment Diabetic Retinopathy Study
GBDT: Gradient boosting decision tree
IDL-CE%: Cholesterol easters to total lipid ratio in intermediate-density lipoprotein particles
LASSO: Logistic regression with the least absolute square shrinkage operator
LR: Logistic regression
PP: Pulse pressure
SEED: The Singapore Epidemiology of Eye Diseases study
UKBB: UK Biobank

## DATA AVAILABILITY

SEED data are available from the Singapore Eye Research Institutional Ethics Committee for researchers who meet the criteria for access to confidential data. Interested researchers can send data access requests to the Singapore Eye Research Institute using the following email address.

The UK Biobank datasets can be requested by bona fide researchers for approved projects, including replication, through **HTTPS://WWW.UKBIOBANK.AC.UK/**.

### Funding Support

This study was supported by the National Medical Research Council, NMRC/StaR/016/2013, NMRC/CIRG/1371/2013, NMRC/CIRG/1417/2015, and OFLCG/001/2017.

### Contribution statement

All authors contributed to the intellectual development of this paper. JL and CS designed the study and supervised data analysis. FH and CNYL wrote the initial draft. FH performed the statistical analyses. SN, CYC, TYW, JL and CS assisted in interpretation of the analyzed data and provided critical corrections of the manuscript. CS is the guarantor of this work and as such had full access to all the data in the study and takes responsibility for the integrity of the data and accuracy of the data analysis. Final version of the paper has been seen and approved by all the authors.

