## Supplementary figures and images for "Machine learning with validation to detect diabetic microvascular complications using clinical and metabolomics data"

### Supplementary Figure S1

**Figure S1. Variable selected by machine learning – metabolites in common**

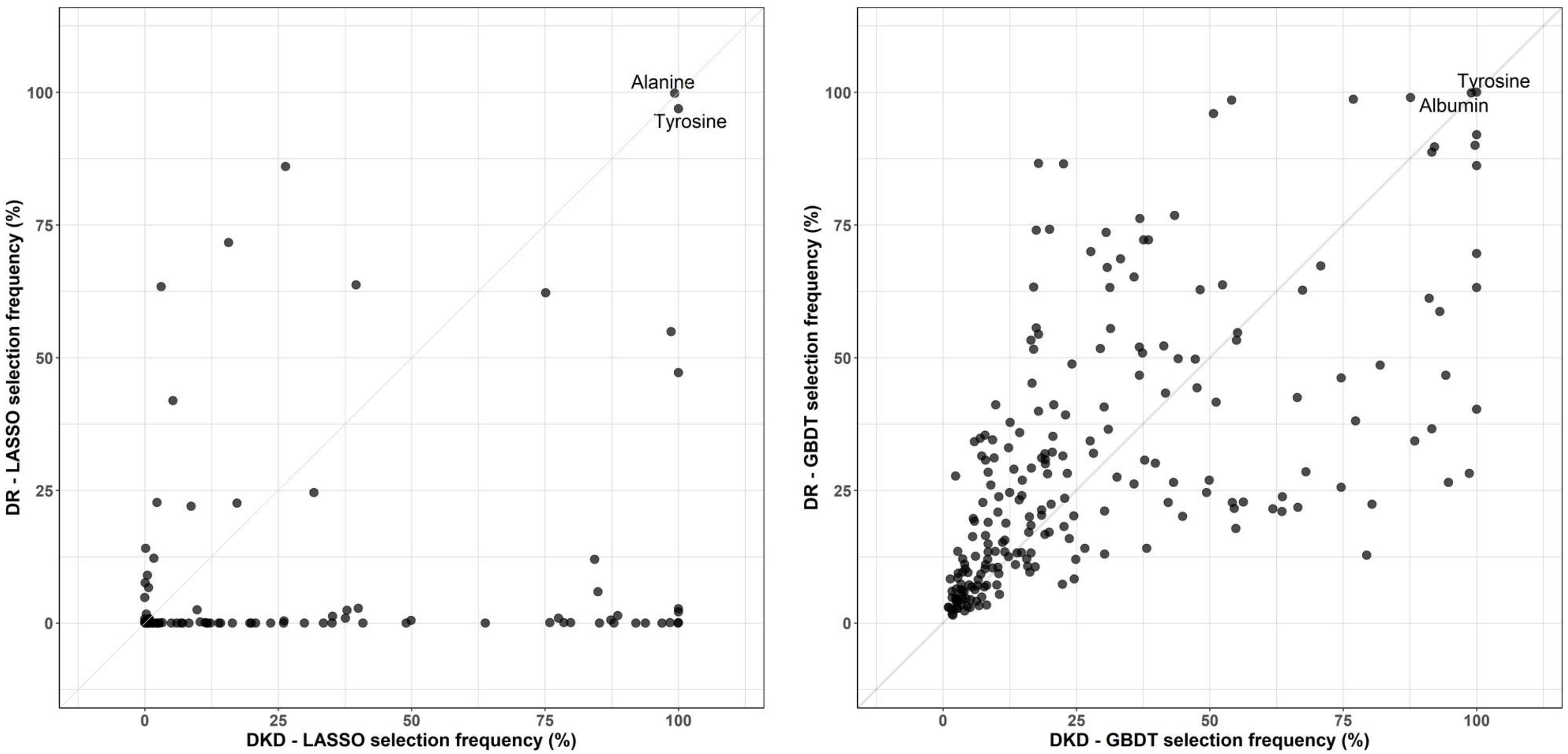

### Supplementary Figure S2

**Figure S2. Variable selection by machine learning – risk factors**

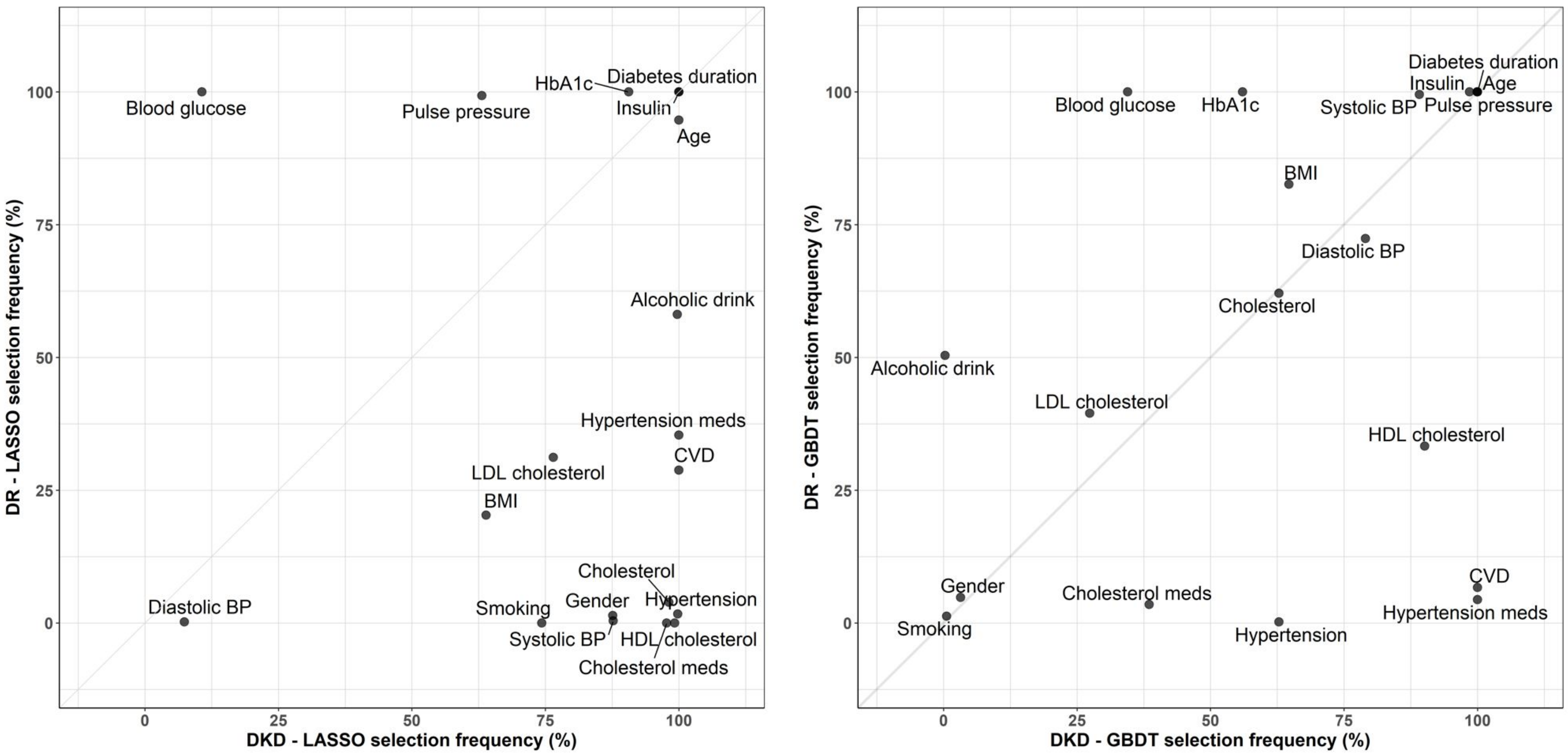

### Supplementary Figure S3

**Figure S3. Bar plots showing the top-50 variables selected by LASSO.**

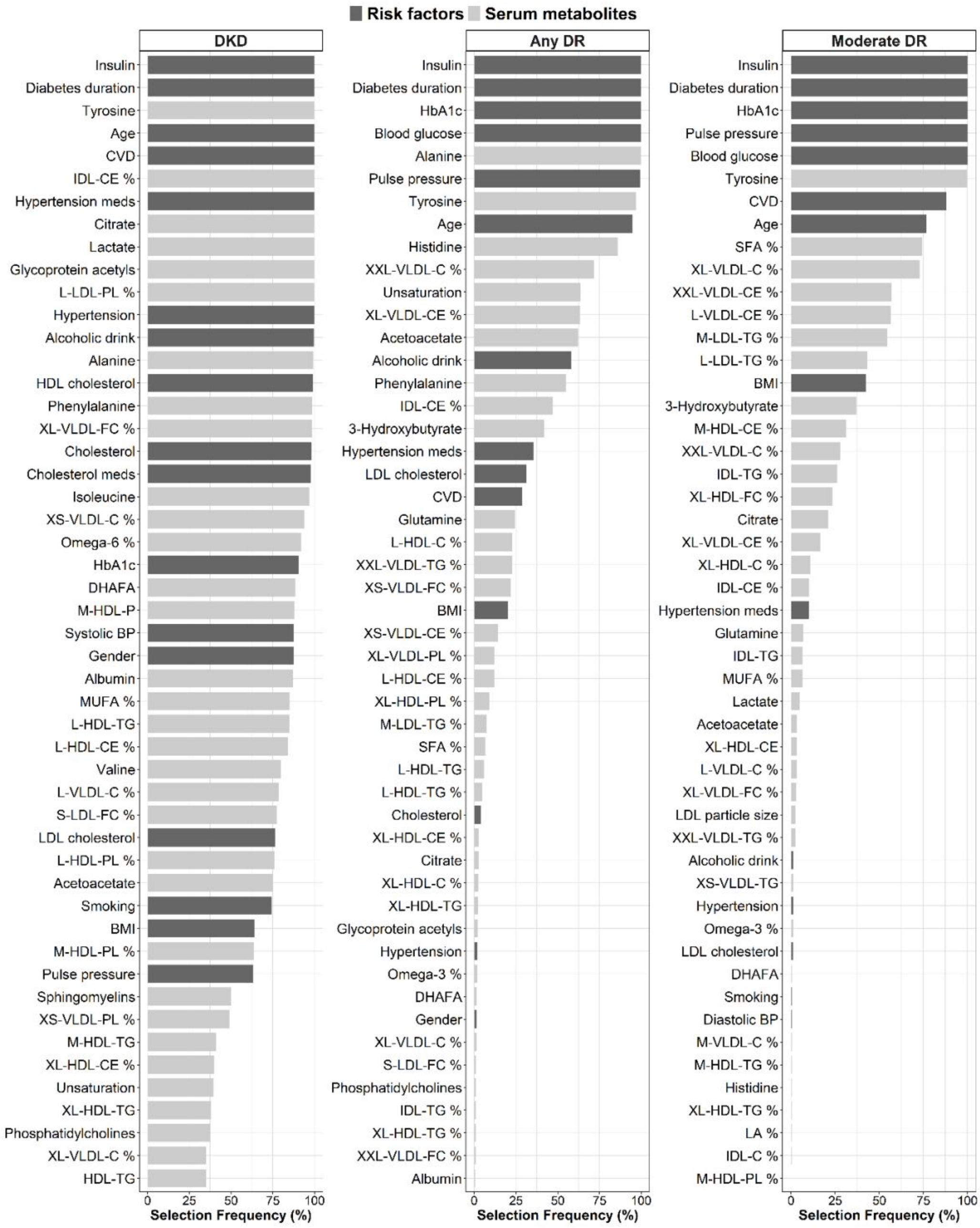

### Supplementary Figure S4

**Figure S4. Bar plots showing the top-50 variables selected by LASSO.**

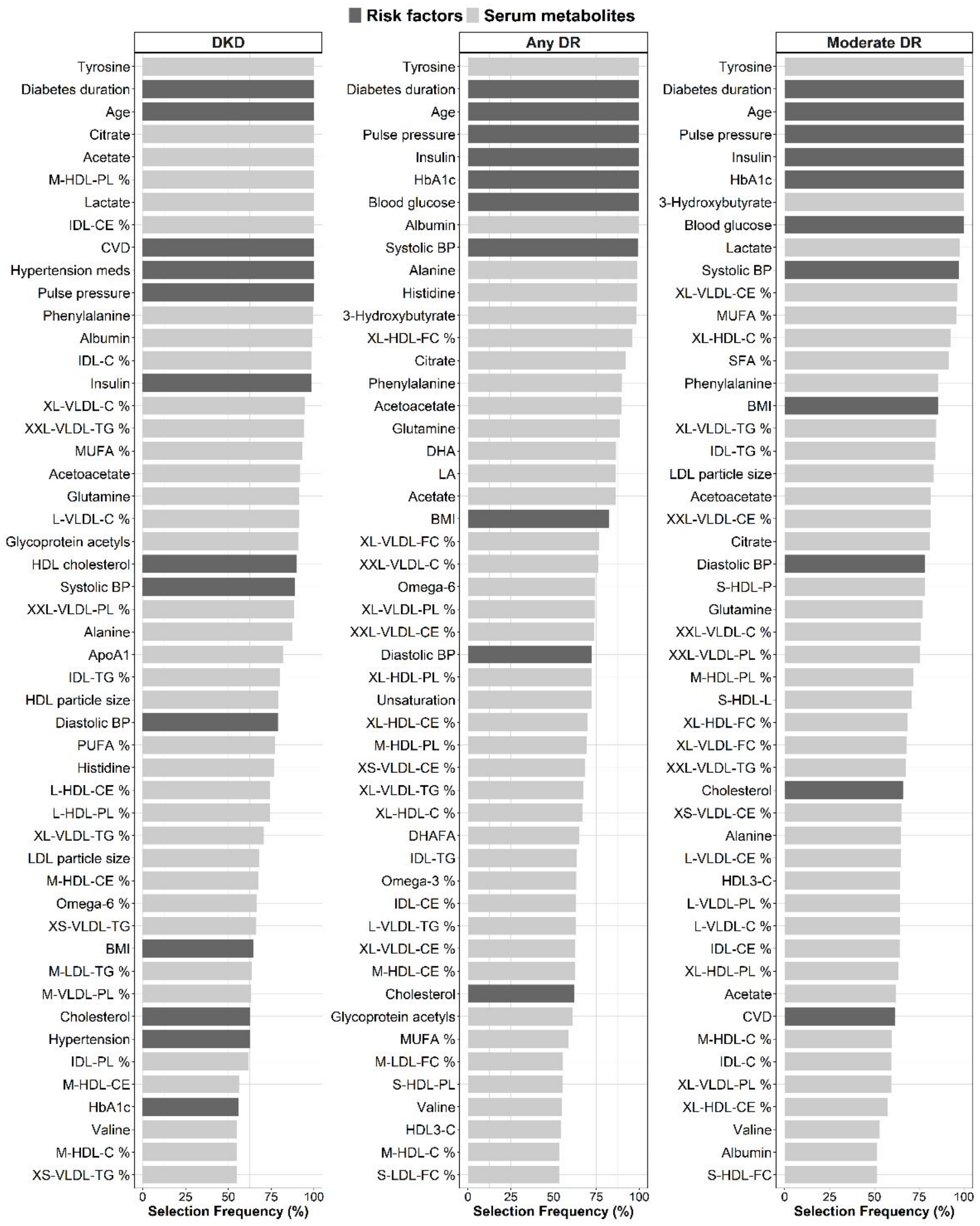
