## Supplementary Table S1 for "Machine learning with validation to detect diabetic microvascular complications using clinical and metabolomics data"

**SUPPLEMENTARY MATERIALS**

**Table S1. List of 239 variables included for machine learning feature selection**

| | | | | | |
|---|---|---|---|---|---|
| **19 risk factors** | | | | | |
| age | alc_cat | anti_chol | anti_ht | bldglu | bmi |
| bpdia_f | bpsys_f | chol | corr_hdlchol | CVD | dbduration |
| gender | hba1c | hypertension | insulin | ldlchol | pulse_press |
| smkcurr | | | | | |
| **220 metabolites** | | | | | |
| AcAce | LA | L-VLDL-P | M-VLDL-L | S-LDL-TG% | XL-VLDL-C% |
| Ace | Lac | L-VLDL-PL | M-VLDL-P | SM | XL-VLDL-CE |
| Ala | LAFA | L-VLDL-PL% | M-VLDL-PL | S-VLDL-C | XL-VLDL-CE% |
| Alb | LDL-D | L-VLDL-TG | M-VLDL-PL% | S-VLDL-C% | XL-VLDL-FC |
| ApoA1 | LDL-TG | L-VLDL-TG% | M-VLDL-TG | S-VLDL-CE | XL-VLDL-FC% |
| ApoB | Leu | M-HDL-C | M-VLDL-TG% | S-VLDL-CE% | XL-VLDL-L |
| ApoBApoA1 | L-HDL-C | M-HDL-C% | PC | S-VLDL-FC | XL-VLDL-P |
| bOHBut | L-HDL-C% | M-HDL-CE | Phe | S-VLDL-FC% | XL-VLDL-PL |
| Cit | L-HDL-CE | M-HDL-CE% | PUFA | S-VLDL-L | XL-VLDL-PL% |
| DHA | L-HDL-CE% | M-HDL-FC | PUFAFA | S-VLDL-P | XL-VLDL-TG |
| DHAFA | L-HDL-FC | M-HDL-FC% | RemnantC | S-VLDL-PL | XL-VLDL-TG% |
| EstC | L-HDL-FC% | M-HDL-L | SerumTG | S-VLDL-PL% | XS-VLDL-C |
| FAw3 | L-HDL-L | M-HDL-P | SFA | S-VLDL-TG | XS-VLDL-C% |
| FAw3FA | L-HDL-P | M-HDL-PL | SFAFA | S-VLDL-TG% | XS-VLDL-CE |
| FAw6 | L-HDL-PL | M-HDL-PL% | S-HDL-C | TGPG | XS-VLDL-CE% |
| FAw6FA | L-HDL-PL% | M-HDL-TG | S-HDL-C% | TotCho | XS-VLDL-FC |
| FreeC | L-HDL-TG | M-HDL-TG% | S-HDL-CE | TotFA | XS-VLDL-FC% |
| Gln | L-HDL-TG% | M-LDL-C | S-HDL-CE% | TotPG | XS-VLDL-L |
| Gp | L-LDL-C | M-LDL-C% | S-HDL-FC | Tyr | XS-VLDL-P |

Confidential

| | | | | | |
|---|---|---|---|---|---|
| HDL-2C | L-LDL-C% | M-LDL-CE | S-HDL-FC% | UnSat | XS-VLDL-PL |
| HDL-3C | L-LDL-CE | M-LDL-CE% | S-HDL-L | Val | XS-VLDL-PL% |
| HDL-D | L-LDL-CE% | M-LDL-FC | S-HDL-P | VLDL-C | XS-VLDL-TG |
| HDL-TG | L-LDL-FC | M-LDL-FC% | S-HDL-PL | VLDL-D | XS-VLDL-TG% |
| His | L-LDL-FC% | M-LDL-L | S-HDL-PL% | VLDL-TG | XXL-VLDL-C |
| IDL-C | L-LDL-L | M-LDL-P | S-HDL-TG | XL-HDL-C | XXL-VLDL-C% |
| IDL-C% | L-LDL-P | M-LDL-PL | S-HDL-TG% | XL-HDL-C% | XXL-VLDL-CE |
| IDL-CE | L-LDL-PL | M-LDL-PL% | S-LDL-C | XL-HDL-CE | XXL-VLDL-CE% |
| IDL-CE% | L-LDL-PL% | M-LDL-TG | S-LDL-C% | XL-HDL-CE% | XXL-VLDL-FC |
| IDL-FC | L-LDL-TG | M-LDL-TG% | S-LDL-CE | XL-HDL-FC | XXL-VLDL-FC% |
| IDL-FC% | L-LDL-TG% | MUFA | S-LDL-CE% | XL-HDL-FC% | XXL-VLDL-L |
| IDL-L | LVLDL-C | MUFAFA | S-LDL-FC | XL-HDL-L | XXL-VLDL-P |
| IDL-P | L-VLDL-C% | M-VLDL-C | S-LDL-FC% | XL-HDL-P | XXL-VLDL-PL |
| IDL-PL | L-VLDL-CE | M-VLDL-C% | S-LDL-L | XL-HDL-PL | XXL-VLDL-PL% |
| IDL-PL% | L-VLDL-CE% | M-VLDL-CE | S-LDL-P | XL-HDL-PL% | XXL-VLDL-TG |
| IDL-TG | L-VLDL-FC | M-VLDL-CE% | S-LDL-PL | XL-HDL-TG | XXL-VLDL-TG% |
| IDL-TG% | L-VLDL-FC% | M-VLDL-FC | S-LDL-PL% | XL-HDL-TG% | |
| Ile | L-VLDL-L | M-VLDL-FC% | S-LDL-TG | XL-VLDL-C | |

*Traditional risk factors: age, gender, systolic blood pressure, hba1c, duration of diabetes, and body mass index. Ethnicity was not adjusted because SEED and UKBB had different ethnicities.

HDL: high-density lipoprotein; IDL: intermediate-density lipoprotein; LDL: low-density lipoprotein; VLDL: very-low-density lipoprotein; L: large; M: medium; S: small; XL: very large, XS: very small; XXL: extremely large; D: mean diameter; C: cholesterol; CE: cholesterol esters; FC: free cholesterol; L: total lipids; P: phospholipids; TG: triglycerides; %: ratio to total lipids; FA: fatty acids;

Confidential
